# Deep learning-Based Correlation Analysis of Pelvic and Spinal Sequences for Enhanced Sagittal Spinal Alignment Prediction

**DOI:** 10.1101/2023.09.17.23295663

**Authors:** Kai Song, Huan Qi, Chi Ma, Pengfei Chi, Junyu Lin, Qiang Yang, Cao Yang, Bing Wang, Fangcai Li, Zezhang Zhu, Weishi Li, Jianguo Zhang, William Lu, Wang Zheng

## Abstract

**Background:** Pelvic Incidence (PI) plays a crucial role in surgical planning. However, it is insufficient for accurately predicting spinal alignment parameters, including Sacral Slope, Pelvic Tilt, and Lumbar Lordosis. We have devised an AI-based method for predicting sagittal spinal alignments with enhanced precision.

**Methods:** We have developed an AI-based system utilizing a Seq2Seq framework to model the spatial correlation between pelvic and spinal key points. This system was trained on a dataset of 337 cases and evaluated using 51 cases obtained from a multi-centre hospital. To address the issue of pelvic rotation, we introduced an Angle Correlation Network. We compared the performance of our AI-based system in predicting spinal alignment against the traditional PI-based method. This comparison was conducted using Mean Absolute Error (MAE) and the Correlation Coefficient (R value) as evaluation metrics.

**Results:** We evaluated the performance of our AI-based system for predicting Sacral Slope (SS), Pelvic Tilt (PT), and Lumbar Lordosis (LL) values. The Pearson correlation coefficient of the AI-based method surpassed that of the PI-based method (0.80 vs 0.67 for SS, 0.73 vs 0.52 for PT, and 0.76 vs 0.48 for LL), indicating a more robust linear relationship between AI predictions and actual values. Additionally, the AI-based method exhibited a lower Mean Absolute Error (MAE) compared to the PI-based method for LL (5.52 vs 6.69), signifying enhanced prediction accuracy.

**Conclusions:** In this study, we demonstrated the potential of an AI-based approach for predicting sagittal spinal alignments with improved precision compared to the traditional PI-based method. The AI-based system, utilizing a Seq2Seq framework and an Angle Correlation Network, exhibited a stronger linear relationship between predicted and actual values for Sacral Slope, Pelvic Tilt, and Lumbar Lordosis, as well as a reduced Mean Absolute Error for Lumbar Lordosis. These findings support the integration of AI in spinal surgery planning and personalized medicine for sagittal alignment evaluation and management.

## Introduction

Bipedalism in Homo sapiens has led to the development of extraordinary sagittal spinopelvic alignments, which enable an energetically economical upright stance due to the reciprocal sagittal curvatures [1]. A healthy, physically mature individual typically possesses a unique spinopelvic alignment characterized by the harmony among pelvic, lumbar, thoracic, and cervical segments. Among these segments, the pelvis lacks movable intervertebral discs and remains relatively constant, even when spinal segments undergo malformation. Consequently, the pelvic sequence can be used to predict original spinal alignments due to its correlation with other spinal segments and invariable morphology [2-17].

Pelvic incidence (PI), defined as the angle between the line perpendicular to the superior plate of S1 at its midpoint and the line connecting this point to the axis of the femoral heads, has been investigated as an invariant morphological parameter of the pelvis [2]. PI has been shown to have strong correlations with pelvic tilt (PT), sacral slope (SS), and lumbar lordosis (LL). As a result, PI is widely used to predict the original spinopelvic alignments of patients with spinal deformity and inform surgical planning [2-17].

However, the current method has several limitations. Firstly, PI is a simplistic graphical angle, providing minimal information about the pelvic sequence and neglecting potentially crucial aspects of pelvic morphology. Secondly, using PI to predict the sagittal sequence of the spine is overly general and lacks specificity, rendering it impossible to discern local spinal sequences. Thirdly, all predictive sequence information is non-visible. Owing to these limitations, the existing method for predicting spinal sagittal alignments is insufficient for surgical design requirements.

Machine learning, particularly deep learning, has demonstrated superiority over human intelligence and has been successfully applied to medical data for clinical purposes [17-20]. Therefore, in this study, we utilized AI technology to conduct correlation analyses of pelvic and spinal sequences, introducing an innovative method for predicting sagittal spinal alignments. Our approach aims to achieve higher accuracy and more direct-viewing information than previous studies.

## Method

### 1. Building an AI-based system to model spatial correlation between pelvic and spinal key points

In this study, we introduce a novel approach for modeling the spatial correlation between pelvic and spinal key points. Our system development involved the inclusion of 337 healthy adults, aged 18 to 35 years, with full-length, free-standing lateral radiographs from our institute. Two orthopedic spine surgeons reached a consensus and annotated the X-ray images by manually recording the spatial coordinates of the following pelvic and spinal key points, as illustrated in Figure 1:

- 12 sacral key points, including posterior edge of the S1 endplate, sacral promontory, posterior edge of the S2-5 superior endplates, anterior edge of the S2-5 superior endplates, posterior edge of the Co1 superior endplate, anterior edge of the Co1 superior endplate, and center of the Co1-Co2 intervertebral junction.
- 11 key points for the Ilium and Ischium, including midpoint of the line connecting the posterior superior iliac spine and posterior inferior iliac spine, apex of the greater sciatic notch, ischial spine, posterior superior edge of the ischial tuberosity, anterior inferior edge of the ischial tuberosity, pubic symphysis, anterior inferior iliac spine, anterior superior iliac spine, line connecting the anterior superior iliac spine and iliac crest apex, and iliac crest apex.
- 2 femoral head centers.
- 59 vertebral body key points for the lumbar, thoracic, and cervical spine.

**Figure 1.**
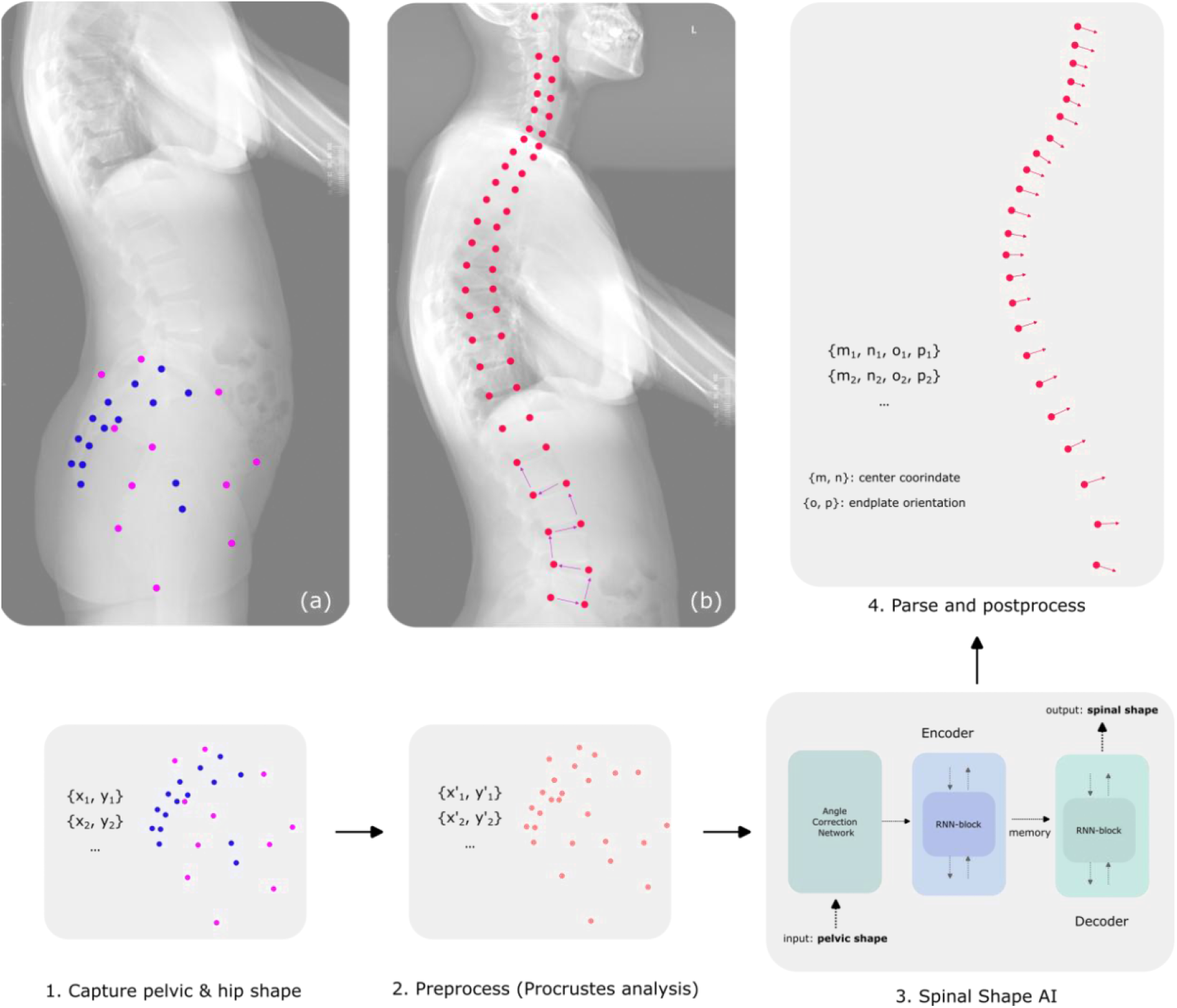
Overview of the proposed AI-based system

The collection of sacrum, femoral head and hip key points is also referred to as “the shape of a pelvis” in the remaining of this paper. The collection of the 59 vertebral body key points is also referred to as “the shape of a spine” in the remaining of this paper. We random selected 63 cases from the 337 cases as a validation dataset for hyperparameter tuning and model selection. The remaining 274 cases were used to train a deep learning model based on the Seq2Seq architecture. We further collected another dataset from multicenter institutes, containing 51 well-positioned full length standing lateral x-ray images, to form a test dataset to evaluate the performance of the proposed system against other established approaches.

Spatial correlation between pelvic and spinal key points were formulated as a supervised regression problem. The independent variable was the shape of a pelvis depicted by a collection of pelvic key point coordinates while the dependent variable was the shape of a spine depicted by a collection of spinal key point coordinates and endplate orientation vectors. A Seq2Seq model is a special type of deep learning models that leverages its recurrent mechanism to model complex long-term spatial or temporal data. Generally, a Seq2Seq model is composed of an encoder and a decoder module. The encoder models the pelvic shape in our case, to capture its intra-class correlation among pelvic key points. The resulting correlation information (or memory) is then fed into the decoder module to model correlation between pelvic shape and spinal shape. In our training setup, the outputs of the decoder module were the regressed offset vectors, forming a zig-zag path, which facilitates learning compared to other formulations Figure 2. The Seq2Seq model was optimized by the Adam optimizer using the combination of a mean-square loss term for coordinate regression and a cosine similarity loss term for orientation regression. A minibatch of 16 samples sufficed in our case to converge after 1,000 epochs.

**Figure 2.**
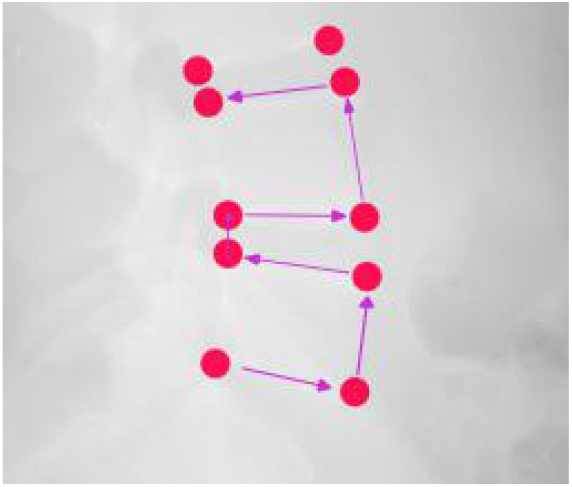
A zig-zag offset scheme to facilitate learning

Rotation is another issue when it comes to exploring pelvic-spinal correspondence. The physiological rotation of pelvic makes it hard to find a “neutral position”. To overcome this issue, we propose a novel Angle Correlation Network (ACN) that rotates input shapes by a predicted angle that would minimize the overall training loss. Throughout the training, ACN learns to address the neutral position problem by itself, as shown in Figure 3.

**Figure 3.**
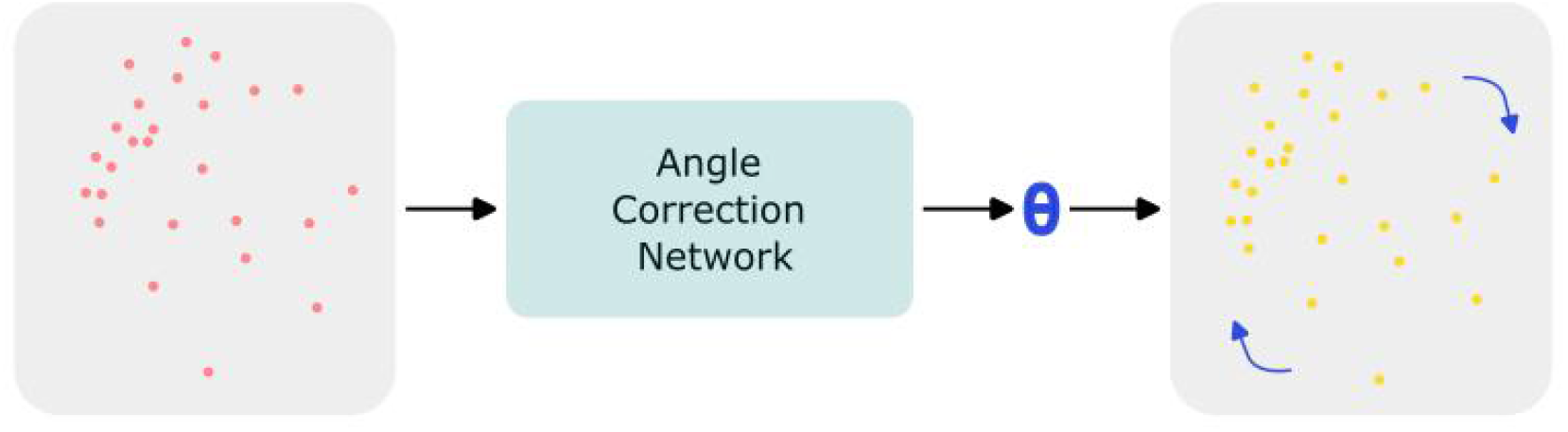
An angle correction network to predict “neutral position.”

During development of the system, we find it critical to conduct pre-processing steps to stabilize training and increase generalization ability. Specifically, a Procrustes analysis was performed on each pair of the pelvic and spinal shape. The aim was to obtain a similar scale, orientation and placement by minimizing shape difference between the training samples so that the model could focus on the given task. The Procrustes analysis contained three steps: translation, uniform scaling, and rotation. The first two steps were straight-forward to perform. The rotation step was conducted by comparing to a randomly selected reference shape from the training dataset. The inverse steps were performed accordingly on the model predictions in the post-processing steps in order to recover the scale and orientation of the input. The overview of the proposed system was shown in Figure 1.

### 2. Performance of the AI-based system compared to a classic clinical study

To evaluate the performance of the AI-based system in predicting sacral slope (SS), pelvic tilt (PT) and lumbar lordosis (LL), a comparison was made with a classic clinical study. The methodology for this comparison is outlined below.

#### Dataset preparation and model selection

The dataset used in this study was divided into three subsets: training, validation, and testing. The training dataset was used to train various AI models, while the validation dataset was employed to fine-tune the models and select the best-performing one based on pre-defined evaluation metrics. The held-out dataset, which contained 51 cases, was reserved for testing the performance of the selected model. A Seq2Seq model was chosen as the AI-based system for this study due to its ability to learn complex patterns and relationships within the data. The best-performing Seq2Seq model was identified according to its performance on the validation dataset.

#### Evaluation metrics

To assess the performance of the selected Seq2Seq model, two evaluation metrics were employed: 1) Mean Absolute Error (MAE): This metric calculates the average magnitude of the errors between the predicted values and the true values of LL and TK. A lower MAE indicates better model performance. 2) Correlation Coefficient (R value): This metric measures the strength and direction of the linear relationship between the predicted values and the true values of SS, PT and LL. A higher R value indicates a strong correlation between the predictions and the true values, suggesting better model performance.

#### Classic clinical study

A classic clinical study (SS=0.63PI+7, PT=0.37PI-7 [3], LL=0.62PI+17.6 [7] was chosen for comparison to evaluate the effectiveness of the AI-based system. In this study, researchers used regression analysis and other statistical methods to predict SS, PT and LL values based on available clinical and radiographic data. The performance of the classic clinical study was assessed using the same evaluation metrics (MAE and R value) as those employed for the AI-based system.

#### Comparison and analysis

We randomly rotate the pelvic images of the 51 test subjects to be tested, with rotation angles ranging from -60 to 60 degrees. After the rotation, we further process the data for comparison. The performance of the AI-based system and the classic clinical study was compared in terms of the evaluation metrics (MAE and R value). The results were then analysed to determine the effectiveness of the AI-based system in predicting SS, PT and LL values relative to the classic clinical study. To ensure a rigorous comparison, potential confounding factors, such as differences in patient populations or data quality, were taken into account. In addition, the generalizability of the AI-based system to diverse patient populations was considered, particularly if the training data was not representative of the target patient group. By comparing the performance of the AI-based system and the classic clinical study using the specified methodology, the study aimed to demonstrate the potential advantages and limitations of using an AI-based approach for predicting SS, PT and LL values in patients.

## Result

The results of our study are presented in Table 1, Table 2, Figure 4, and Figure 5. Table 1 presents the details of the 51 testing cases, and Table 2 presents the statistical description, Mean Absolute Error (MAE) and Pearson correlation coefficient for PI and AI predictions on SS, PT, and LL variables.

**Table 1:** Performance of the testing cases.

| Case | Pelvic | PI | PI prediction |  |  | AI prediction<br>(Seq2Seq-based) |  |  | Ture Value |  |  |
| --- | --- | --- | --- | --- | --- | --- | --- | --- | --- | --- | --- |
|  | Random<br>rotation |  | SS① | PT② | LL③ | SS | PT | LL | SS | PT | LL |
| 1 | 6.7 | 42.7 | 33.9 | 9.2 | 44.1 | 27.5 | 15.2 | 40.6 | 23.8 | 18.9 | 33.8 |
| 2 | -33.5 | 49.2 | 38.0 | 11.7 | 48.1 | 43.7 | 5.5 | 56.8 | 42.9 | 6.3 | 54.0 |
| 3 | 3.9 | 48.4 | 37.5 | 11.4 | 47.6 | 36.2 | 12.3 | 49.2 | 36.7 | 11.7 | 55.7 |
| 4 | -1.6 | 41.5 | 33.2 | 8.8 | 43.3 | 31.8 | 9.7 | 44.9 | 31.1 | 10.5 | 37.6 |
| 5 | -8.4 | 50.3 | 38.7 | 12.1 | 48.8 | 38.2 | 12.0 | 51.3 | 44.1 | 6.2 | 57.2 |
| 6 | -30.5 | 42.2 | 33.6 | 9.1 | 43.8 | 36.8 | 5.5 | 49.9 | 36.6 | 5.7 | 48.5 |
| 7 | 47.3 | 56.1 | 42.3 | 14.3 | 52.4 | 44.7 | 11.4 | 57.7 | 44.4 | 11.7 | 52.4 |
| 8 | -3.4 | 38.6 | 31.3 | 7.7 | 41.5 | 33.8 | 4.8 | 47.0 | 30.4 | 8.3 | 49.6 |
| 9 | 44.4 | 48.3 | 37.4 | 11.4 | 47.5 | 25.4 | 22.9 | 38.5 | 27.9 | 20.4 | 21.8 |
| 10 | -18.9 | 52.7 | 40.2 | 13.0 | 50.3 | 39.9 | 12.8 | 52.9 | 42.4 | 10.4 | 55.0 |
| 11 | 18.4 | 51.6 | 39.5 | 12.6 | 49.6 | 39.5 | 12.1 | 52.5 | 38.9 | 12.7 | 46.5 |
| 12 | -46.8 | 59.6 | 44.6 | 15.7 | 54.6 | 46.5 | 13.2 | 59.6 | 53.1 | 6.5 | 65.2 |
| 13 | 12.5 | 42.6 | 33.9 | 9.2 | 44.0 | 33.2 | 9.4 | 46.4 | 28.9 | 13.8 | 40.9 |
| 14 | -2.4 | 36.1 | 29.7 | 6.7 | 40.0 | 29.4 | 6.7 | 42.5 | 25.8 | 10.3 | 31.3 |
| 15 | 9.6 | 43.6 | 34.5 | 9.6 | 44.6 | 32.9 | 10.7 | 45.9 | 39.2 | 4.4 | 47.7 |
| 16 | 18.7 | 42.0 | 33.5 | 9.0 | 43.6 | 30.7 | 11.3 | 43.8 | 35.3 | 6.7 | 47.1 |
| 17 | 48.5 | 53.5 | 40.7 | 13.3 | 50.8 | 33.9 | 19.6 | 47.0 | 29.6 | 23.9 | 39.9 |
| 18 | -3.9 | 42.1 | 33.5 | 9.0 | 43.7 | 37.0 | 5.1 | 50.1 | 30.4 | 11.6 | 41.9 |
| 19 | 14.9 | 43.7 | 34.5 | 9.6 | 44.7 | 44.8 | -1.1 | 58.0 | 41.5 | 2.2 | 51.5 |
| 20 | 9.6 | 43.4 | 34.3 | 9.5 | 44.5 | 37.3 | 6.1 | 50.4 | 33.0 | 10.4 | 40.3 |
| 21 | -11.8 | 44.6 | 35.1 | 10.0 | 45.3 | 39.0 | 5.7 | 52.1 | 35.3 | 9.3 | 47.4 |
| 22 | 54.3 | 47.5 | 36.9 | 11.0 | 47.1 | 35.5 | 12.0 | 48.6 | 32.9 | 14.6 | 31.7 |
| 23 | 8.0 | 65.2 | 48.1 | 17.8 | 58.0 | 51.1 | 14.2 | 64.1 | 47.6 | 17.6 | 56.2 |
| 24 | -15.3 | 52.7 | 40.2 | 13.0 | 50.3 | 49.2 | 3.5 | 62.4 | 45.4 | 7.3 | 59.2 |
| 25 | 54.8 | 62.5 | 46.4 | 16.8 | 56.4 | 47.1 | 15.4 | 60.1 | 49.5 | 13.0 | 57.2 |
| 26 | -6.7 | 38.0 | 31.0 | 7.4 | 41.2 | 35.6 | 2.4 | 48.7 | 33.7 | 4.3 | 47.9 |
| 27 | -16.6 | 55.5 | 42.0 | 14.1 | 52.0 | 43.7 | 11.8 | 56.7 | 44.5 | 11.0 | 48.0 |
| 28 | 55.8 | 47.4 | 36.8 | 11.0 | 47.0 | 23.0 | 24.3 | 36.1 | 34.1 | 13.3 | 48.3 |
| 29 | 33.6 | 34.0 | 28.4 | 5.9 | 38.7 | 25.6 | 8.4 | 38.8 | 26.6 | 7.4 | 34.5 |
| 30 | -3.4 | 43.3 | 34.3 | 9.5 | 44.4 | 51.5 | -8.2 | 64.6 | 45.4 | -2.1 | 64.9 |
| 31 | -36.8 | 34.2 | 28.6 | 6.0 | 38.8 | 31.5 | 2.8 | 44.6 | 24.2 | 10.1 | 38.9 |
| 32 | 7.6 | 41.9 | 33.4 | 8.9 | 43.6 | 33.3 | 8.6 | 46.4 | 38.5 | 3.3 | 49.1 |
| 33 | 55.0 | 36.9 | 30.2 | 7.0 | 40.5 | 14.8 | 22.1 | 27.8 | 19.7 | 17.2 | 27.1 |
| 34 | 56.4 | 39.6 | 31.9 | 8.0 | 42.2 | 26.8 | 12.8 | 40.0 | 32.2 | 7.4 | 42.9 |
| 35 | -38.5 | 53.5 | 40.7 | 13.3 | 50.8 | 39.7 | 13.7 | 52.8 | 30.9 | 22.6 | 37.1 |
| 36 | 19.9 | 41.7 | 33.2 | 8.8 | 43.5 | 38.5 | 3.2 | 51.6 | 44.5 | -2.8 | 54.9 |
| 37 | -54.0 | 65.8 | 48.4 | 18.0 | 58.4 | 52.5 | 13.3 | 65.5 | 44.2 | 21.6 | 59.1 |
| 38 | -23.4 | 42.8 | 34.0 | 9.3 | 44.1 | 46.4 | -3.5 | 59.4 | 35.4 | 7.4 | 58.7 |
| 39 | -22.9 | 45.6 | 35.7 | 10.3 | 45.9 | 47.9 | -2.3 | 61.0 | 46.0 | -0.4 | 56.4 |
| 40 | 46.4 | 35.9 | 29.6 | 6.6 | 39.9 | 32.9 | 3.0 | 46.0 | 32.0 | 3.9 | 48.1 |
| 41 | -47.1 | 41.4 | 33.1 | 8.7 | 43.3 | 38.5 | 2.9 | 51.7 | 33.5 | 7.9 | 43.2 |
| 42 | -1.1 | 51.7 | 39.6 | 12.6 | 49.7 | 42.3 | 9.4 | 55.3 | 38.7 | 13.0 | 56.1 |
| 43 | -23.0 | 63.4 | 47.0 | 17.1 | 56.9 | 36.6 | 26.8 | 49.7 | 41.3 | 22.2 | 54.0 |
| 44 | 35.5 | 39.7 | 32.0 | 8.1 | 42.2 | 26.2 | 13.5 | 39.2 | 34.8 | 4.9 | 42.3 |
| 45 | 22.7 | 57.1 | 43.0 | 14.7 | 53.0 | 49.5 | 7.6 | 62.6 | 44.4 | 12.7 | 69.9 |
| 46 | 53.0 | 44.1 | 34.8 | 9.8 | 44.9 | 34.5 | 9.6 | 47.7 | 39.1 | 5.0 | 58.5 |
| 47 | 43.8 | 50.2 | 38.6 | 12.1 | 48.7 | 34.9 | 15.3 | 48.0 | 37.1 | 13.1 | 45.1 |
| 48 | -6.0 | 36.6 | 30.1 | 6.9 | 40.3 | 32.9 | 3.7 | 46.1 | 31.2 | 5.4 | 52.2 |
| 49 | 32.5 | 34.6 | 28.8 | 6.1 | 39.1 | 24.5 | 10.1 | 37.6 | 33.2 | 1.3 | 39.4 |
| 50 | 0.7 | 44.2 | 34.8 | 9.8 | 45.0 | 39.0 | 5.2 | 52.2 | 39.6 | 4.5 | 48.4 |
| 51 | -37.1 | 47.7 | 37.0 | 11.1 | 47.2 | 31.7 | 16.0 | 44.8 | 30.1 | 17.6 | 47.1 |

① $SS=0.63PI+7$ [3]
② $PT=0.37PI-7$ [3]
③ $LL=0.62PI+17.6$ [7]

**Table 2:** Performance Comparison of PI and AI Predictions in Random Rotation.

|  | True values | Random rotation -60°to +60° |  |  |  |  |  |
| --- | --- | --- | --- | --- | --- | --- | --- |
|  |  | PI prediction (① ② ③) |  |  | AI prediction (Seq2Seq-based) |  |  |
| | $x \pm s$ | $x \pm s$ | R④ | MAE | $x \pm s$ | R④ | MAE |
| PI | $46.41 \pm 8.08$ | - | - | - | - | - | - |
| SS | $36.49 \pm 7.28$ | $36.24 \pm 5.14$ | 0.67 | 4.24 | $36.85 \pm 8.17$ | 0.80 | 4.08 |
| PT | $9.92 \pm 6.32$ | $10.64 \pm 3.10$ | 0.52 | 4.28 | $9.57 \pm 6.93$ | 0.73 | 4.08 |
| LL | $47.87 \pm 9.93$ | $46.4 \pm 5.1$ | 0.48 | 6.69 | $49.95 \pm 8.17$ | 0.76 | 5.51 |

① $SS=0.63PI+7$ [3]
② $PT=0.37PI-7$ [3]
③ $LL=0.62PI+17.6$ [7]
④ Pearson correlation coefficient with true value

**Figure 4.**
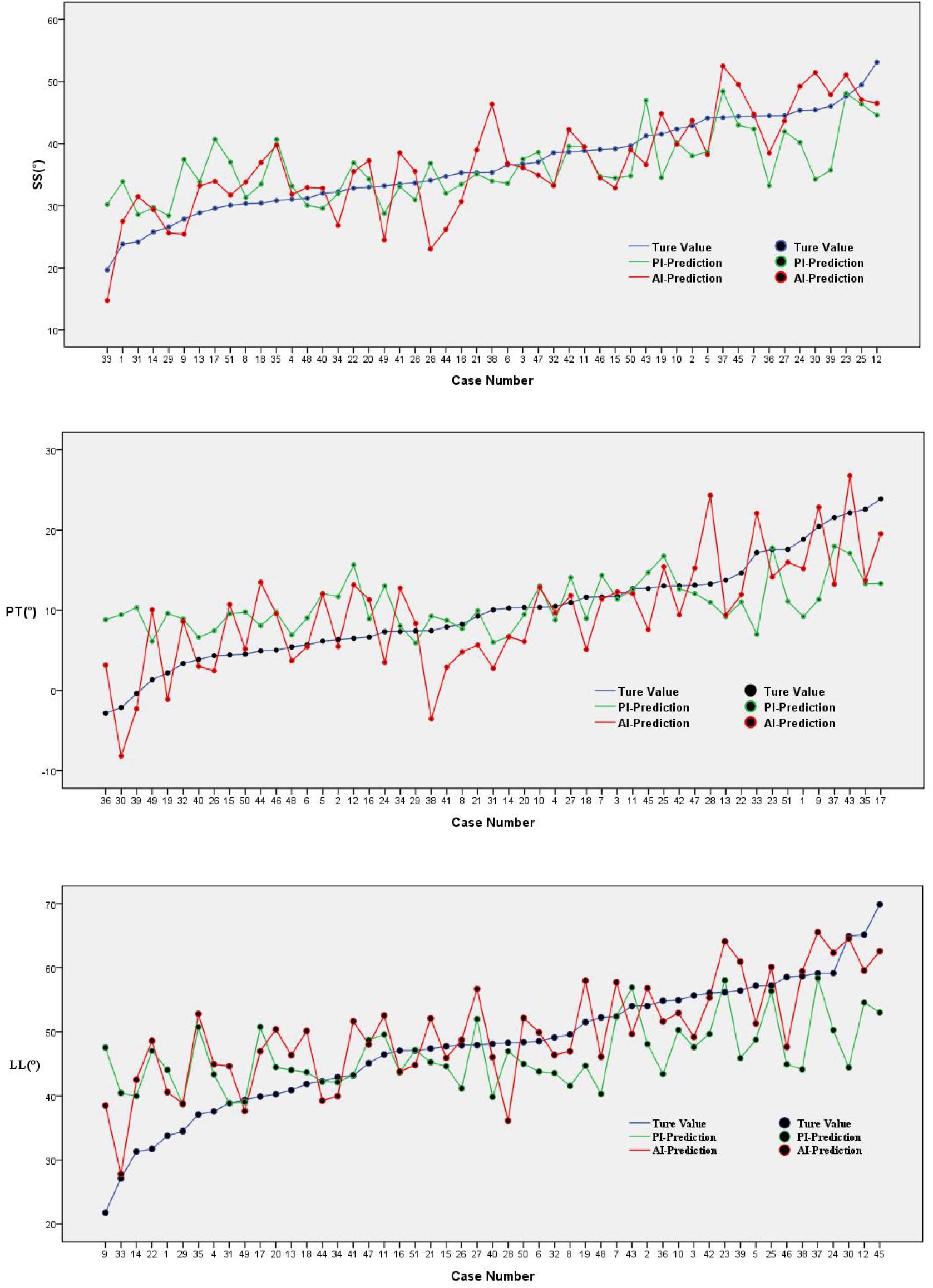
Comparison of True Values, PI Predictions, and AI Predictions for SS, PT, and LL Variables

**Figure 5.**
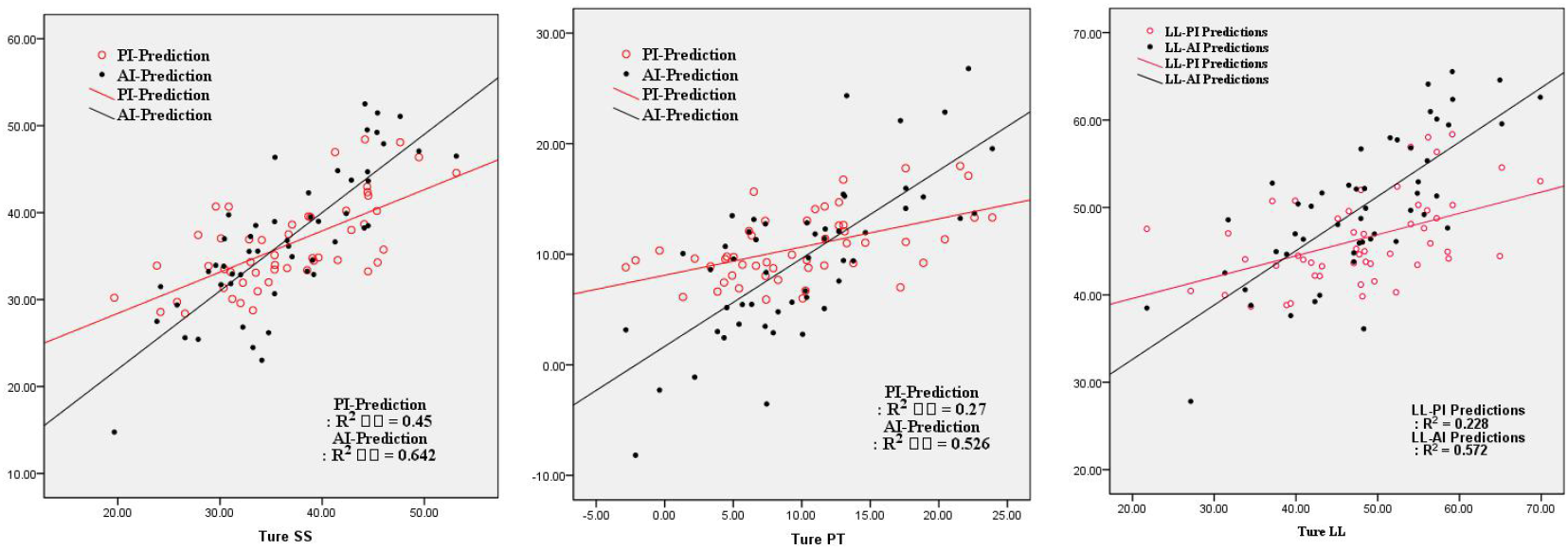
Comparison of MAE and Pearson Correlation for True Values, PI Predictions, and AI Predictions of SS, PT, and LL Variables

For the SS, PT and LL variable, the Pearson correlation coefficient for AI predictions was higher than that for PI predictions (0.80 vs 0.67, 0.73 vs 0.52, 0.76 vs 0.48), demonstrating a stronger linear relationship between AI predictions and the true values. In addition, AI predictions showed a lower MAE compared to PI predictions on LL (5.52 vs 6.69).

From Figures 4 and 5, we can see that the PI predictions for SS, PT, and LL perform reasonably well in the median range but are less satisfactory at the two ends of the normal distribution. In comparison, AI not only performs well in the median range but also demonstrates impressive results at the extreme values.

The AI prediction method consistently outperforms the PI prediction method.

## Discussion

In the 1990s, Legaye and Dubousset introduced sagittal pelvic parameters (PI, PT, and SS) to the field of spinal research [2]. Lots of scholars used these pelvic parameters, along with lumbar lordosis (LL) and other factors, to analyze and classify the sagittal sequence of normal individuals in an upright posture [2-17]. Their work revealed the relationship between these parameters and numerous spinal disorders.

More importantly, many studies have confirmed that having a good match between the pelvic parameters and the lumbar sequence is crucial for diagnosing and treating spinal disorders, as well as for avoiding adverse postoperative events. As a result, the evaluation systems for pelvic parameters (PI, SS and PT) and LL have become a research hotspot in the spinal field.

Vialle, R et al recruited 300 healthy volunteers and abstained SS=0.63PI+7 and PT=0.37PI-7 [3]. Schwab et al. proposed that LL= PI±10° can be used as a rough approximation of the relationship between pelvic parameters and lumbar lordosis in an upright posture [10]. Mac-Thiong and Roussouly, on the other hand, derived a more accurate formula based on a larger sample size: LL=0.62PI+17.6 [7].

Researchers from various countries have also developed formulas for predicting LL using PI as the basis, based on their own experimental samples. In the study, due to the test sample size (51 cases), R-value was not as high as in previous literature, but the result showed that AI was better than PI in predicting SS, PT and LL.

To date, all mainstream spinal-pelvic sagittal studies both domestically and internationally have been based solely on the PI system [2-17]. However, PI is just a simple angular measurement of pelvic morphology. Although it largely reflects the corresponding pelvic morphology, a single angle cannot accurately represent the many specific features of an individual’s pelvic morphology. Different features of pelvic morphology may have varying impacts on spinal matching. Therefore, relying solely on the PI system to predict LL using pelvic parameters may cause us to overlook other useful pelvic morphological parameters.

We cannot deny that in the era of human computational power, PI has been the best and most straightforward pelvic morphological parameter for predicting LL. However, we have now entered the era of AI computational power, where AI’s superior computing capabilities eliminate the need to consider the significant costs of human labor and time [20]. Therefore, through AI-based analysis and prediction of the pelvis and spine, we can avoid losing any data and not waste any potential meaningful opportunities [19]. We developed an AI-based system using a Seq2Seq algorithm to model the spatial correlation between pelvic and spinal keypoints [18]. The system was trained on 274 cases and evaluated on a test dataset of 65 cases. A novel Angle Correlation Network was proposed to address the issue of rotation. The system outperformed a classic clinical study in terms of both mean absolute error and correlation coefficient, using MAE and R value as evaluation metric. In this research, we manually annotated key points of the pelvis and spine on standing lateral full-length radiographs. The relationships between these key points generate much more data than the simple relationship between PI and LL. In fact, not only can we obtain the LL value of the spinal sequence, but we can also obtain more detailed local sequence relationships, such as the lower lumbar lordosis angle of L4-S1, and so on. All this data can be presented in a visualized form, which is something the PI system cannot achieve.

There are still some shortcomings in this study, as we manually annotated key points of the pelvis and predicted their relationships. Further research is needed to: 1) improve AI-based automatic recognition of key points and prediction, 2) abandon key point recognition and use purely morphological learning for prediction, which relies on a more extensive data sample, and 3) improve three-dimensional morphological recognition and prediction using technology like CT three-dimensional reconstruction, EOS, and others. We firmly believe that this is just the beginning of AI-based research on the sagittal sequence of the spine and pelvis, and it will ultimately far surpass human capabilities.

## Conclusion

In conclusion, this study demonstrates the potential of deep learning-based methods in predicting spinal sagittal alignments. Our Seq2Seq-based AI model outperformed the traditional method based on PI. By providing accurate and detailed predictions, the AI model has a potential to improve surgical planning and outcomes for patients with spinal diseases.

## Data Availability

All data produced in the present study are available upon reasonable request to the authors

## References

1. Lovejoy, C. O. (2005). The natural history of human gait and posture. Part 1. Spine and pelvis. Gait & Posture, 21(1), 95–112.

2. Legaye, J., Duval-Beaupère, G., Hecquet, J., & Marty, C. (1998). Pelvic incidence: a fundamental pelvic parameter for three-dimensional regulation of spinal sagittal curves. European Spine Journal, 7(2), 99–103.

3. Vialle, R., Levassor, N., Rillardon, L., Templier, A., Skalli, W., & Guigui, P. (2005). Radiographic Analysis of the Sagittal Alignment and Balance of the Spine in Asymptomatic Subjects. The Journal of Bone and Joint Surgery. American volume, 87(2), 260–267. doi:10.2106/JBJS.D.02043

4. Roussouly, P., Gollogly, S., Berthonnaud, E., & Dimnet, J. (2005). Classification of the normal variation in the sagittal alignment of the human lumbar spine and pelvis in the standing position. European Spine Journal, 16(3), 227–234. DOI: 10.1007/s00586-005-0013-8

5. Boulay, C., Tardieu, C., Hecquet, J., Benaim, C., Mouilleseaux, B., Marty, C., … & Skalli, W. (2006). Sagittal alignment of spine and pelvis regulated by pelvic incidence: standard values and prediction of lordosis. European Spine Journal, 15(4), 415–422. DOI: 10.1007/s00586-005-0984-5

6. Barrey, C., Jund, J., Noseda, O., & Roussouly, P. (2007). Sagittal balance of the pelvis-spine complex and lumbar degenerative diseases. A comparative study about 85 cases. European Spine Journal, 16(9), 1459–1467. DOI: 10.1007/s00586-006-0294-6

7. Mac-Thiong, J. M., Labelle, H., Charlebois, M., Huot, M. P., & de Guise, J. A. (2007). Sagittal spinopelvic balance in normal children and adolescents. European Spine Journal, 16(2), 227–234. DOI: 10.1007/s00586-005-0013-8

8. Barrey, C., Jund, J., Noseda, O., & Roussouly, P. (2007). Sagittal balance of the pelvis-spine complex and lumbar degenerative diseases. A comparative study about 85 cases. European Spine Journal, 16(9), 1459–1467.

9. Schwab, F., Lafage, V., Patel, A., & Farcy, J. P. (2009). Sagittal plane considerations and the pelvis in the adult patient. Spine, 34(17), 1828–1833. DOI: 10.1097/BRS.0b013e3181a13c08

10. Schwab, F., Patel, A., Ungar, B., Farcy, J. P., & Lafage, V. (2010). Adult spinal deformity-postoperative standing imbalance: how much can you tolerate? An overview of key parameters in assessing alignment and planning corrective surgery. Spine, 35(25), 2224–2231. DOI: 10.1097/BRS.0b013e3181ee6bd4

11. Roussouly, P., & Nnadi, C. (2010). Sagittal plane deformity: an overview of interpretation and management. European Spine Journal, 19(11), 1824–1836.

12. Mac-Thiong, J. M., Roussouly, P., Berthonnaud, E., & Guigui, P. (2011). Age- and sex-related variations in sagittal sacropelvic morphology and balance in asymptomatic adults. European Spine Journal, 20(Suppl 5), 572–577.

13. Roussouly, P., & Pinheiro-Franco, J. L. (2011). Biomechanical analysis of the spino-pelvic organization and adaptation in pathology. European Spine Journal, 20(Suppl 5), 609–618.

14. Roussouly, P., & Pinheiro-Franco, J. L. (2011). Biomechanical analysis of the spino-pelvic organization and adaptation in pathology. European Spine Journal, 20(Suppl 5), 609–618. DOI: 10.1007/s00586-011-1928-x

15. Protopsaltis, T., Schwab, F., Bronsard, N., Smith, J. S., Klineberg, E., Mundis, G., & Ames, C. P. (2014). TheT1 pelvic angle, a novel radiographic measure of global sagittal deformity, accounts for both spinal inclination and pelvic tilt and correlates with health-related quality of life. Journal of Bone and Joint Surgery, 96(19), 1631–1640.

16. Zhu, Z., Xu, L., Zhu, F., Jiang, L., Wang, Z., Liu, Z., … & Qian, B. (2014). Sagittal alignment of spine and pelvis in asymptomatic adults: norms in Chinese populations. Spine, 39(1), E1–E6. DOI: 10.1097/BRS.0000000000000072

17. Laouissat, F., Sebaaly, A., Gehrchen, M., & Roussouly, P. (2018). Classification of normal sagittal spine alignment: refounding the Roussouly classification. European Spine Journal, 27(8), 2001–2011.

18. Sutskever, I., Vinyals, O., & Le, Q. V. (2014). Sequence to sequence learning with neural networks. Advances in Neural Information Processing Systems, 27, 3104–3112.

19. LeCun, Y., Bengio, Y., & Hinton, G. (2015). Deep learning. Nature, 521(7553), 436–444. DOI: 10.1038/nature14539

20. Goodfellow, I., Bengio, Y., & Courville, A. (2016). Deep learning. MIT Press.

